# Maternal immunity, cesarean delivery, and childhood neuropsychiatric risk in 1.18 million births

**DOI:** 10.64898/2026.05.27.26354231

**Authors:** Benjamin Kramer, Steven A. Kushner, Andrey Rzhetsky

## Abstract

Maternal infection, immune disease, and delivery mode are plausible influences on early brain development. We analyzed 1,179,611 US Merative MarketScan mother-child pairs (2003–2024), including 259,339 non-twin siblings in 123,926 families. Population models screened 18 perinatal exposures against 13 childhood psychiatric/neurodevelopmental diagnosis-count outcomes; sibling fixed effects tested robustness to stable family-level confounding. Cesarean delivery was associated with higher composite neurodevelopmental diagnosis counts in pairs (23.4%) and siblings (25.0%) and with ADHD in siblings (38.8%; FDR *q* = 0.025). Autism was elevated in pairs (20.0%) but not supported within families (5.0%; *p* = 0.87). Claims-defined no-labor/no-repeat cesarean showed stronger lower-risk-birth associations for composite neurodevelopmental burden (48.0%), autism (44.9%), speech/language disorders (41.0%), and ADHD (24.1%). Maternal infection/immune-mediated disease, preterm birth, and advanced maternal age were additional population signals.

---

Fetal development and birth are short epochs in human development that have a major influence on future health. Maternal infection or immune-mediated disease and associated medications, maternal age, preterm birth, and mode of delivery contribute to early developmental biology ^1–8^, but their impacts on childhood neurodevelopmental and psychiatric disorders are difficult to quantify. A conventional population-level cohort can detect small-effect associations, but it could also be biased by confounding family-specific genetic, household, obstetric, and healthcare-access factors. A sibling design suffers from a smaller sample size, but accounts for the complex family-specific covariates that allow testing whether exposed children differ from their unexposed siblings born to the same mother ^9–12^.

Cesarean delivery (CD) is the clearest example of this dilemma of analysis design. CD accounts for more than one-third of births in the US^13^ and has been reported to be associated with autism spectrum disorders (ASDs), attention deficit hyperactivity disorder (ADHD), and other broader psychiatric outcomes ^14–17^. CD has also been reported to be associated with immune-mediated, respiratory, infection-related, and metabolic childhood outcomes^18–23^. However, delivery mode is also affected by pregnancy complications, parental risks, household context, and access and use of healthcare. Studies of Swedish siblings questioned the apparent population-level associations of CD-ASDs and CD-ADHD^14,15^. Genetically sensitive designs remain the key missing step in such analyses ^16,17^.

The question is therefore broader than whether CD predicts ASDs. We designed the study as a biologically motivated perinatal-exposure atlas. We ask whether maternal infection or immunity-related disease during pregnancy, delivery mode, preterm birth, birth weight, and maternal age are associated with childhood-onset neurodevelopmental and psychiatric diagnoses; and then, where siblings’ data allow, whether those associations persist after accounting for stable family-specific factors. We also separated cesarean delivery into mutually exclusive priority-coded claims categories: no-labor/no-repeat, repeat/prior-cesarean without detected labor-onset, and labor-onset cesarean. This decomposition tested whether the signal strengthened for the subtype closest to elective or pre-labor cesarean.

We analyzed 1,179,611 US mother-child pairs from Merative MarketScan Commercial Claims (2003 to 2024), including a nested non-twin sibling cohort of 259,339 children in 123,926 families. The pair cohort maximized statistical power to detect associations of 18 perinatal exposures and 13 psychiatric/neurodevelopmental outcomes. The sibling cohort controlled for time-invariant family-specific confounding through mother fixed effects. Our goal was to identify which pair-cohort-derived signals remain robust under the more conservative sibling design. CD became the focus of downstream subtype and sensitivity analyses because it was a common, potentially modifiable delivery-mode exposure with family-controlled support, whereas preterm birth and low birth weight were stronger but well-established and less clinically elective risk factors.

## Results

### Cohort characteristics

The analytical cohort comprised 1,179,611 mother-newborn pairs with a mean observation after birth of 3.43 years (median 2.35, IQR 1.24 to 4.55). The prevalence of cesarean delivery was 36.7% (claims-defined no-labor/no-repeat cesarean 10.7%, repeat/prior-cesarean without detected labor-onset 16.2%, labor-onset cesarean 9.8%), preterm birth 7.1%, and low birth weight 4.0%. The non-twin sibling subcohort comprised 259,339 children in 123,926 families after excluding 16,986 twins and 999 singleton remnants created by twin removal; all exposure standardized mean differences between the full and sibling cohorts were ≤ 0.11 (Table 1; Extended Data Tables 1 and 2).

**Table 1:** Cohort characteristics for the population and sibling fixed-effects analyses. Counts and proportions are shown for the full analytic cohort and the non-twin sibling subcohort. The compact main table lists the design-critical cohort features; fuller exposure balance and follow-up distributions are provided in Extended Data Tables 1 and 2.

| Characteristic | Full cohort | Sibling subcohort |
| --- | --- | --- |
| Children | 1,179,611 | 259,339 |
| Families | – | 123,926 |
| Person-years of follow-up | 4,041,343 | 1,219,429 |
| Mean follow-up, years | 3.43 | 4.70 |
| Median follow-up, years | 2.35 | 3.97 |
| Cesarean delivery, any | 433,171 (36.7%) | 81,705 (31.5%) |
| Claims-defined no-labor/no-repeat cesarean | 125,665 (10.7%) | 20,346 (7.8%) |
| Repeat/prior-cesarean without detected labor-onset | 191,595 (16.2%) | 40,778 (15.7%) |
| Claims-defined labor-onset cesarean | 115,911 (9.8%) | 20,581 (7.9%) |
| Cesarean-discordant families in primary sibling support set | – | 13,189 (10.6%) |
| Preterm birth (<37 weeks) | 84,291 (7.1%) | 13,866 (5.3%) |
| Low birth weight (<2500 g) | 46,978 (4.0%) | 6,755 (2.6%) |
| Female | 574,819 (48.7%) | 125,753 (48.5%) |

The main displays used the final 13-outcome DSM-5 psychiatric and neurodevelopmental panel. Intellectual disability was defined but excluded from population models because theta-boundary non-convergence prevented stable negative-binomial estimation (641 affected children; 4,880 diagnosis-days); it was retained only in converged sibling fixed-effects matrices.

### The pairs cohort identifies a broad perinatal risk pattern

Analysis of 234 exposure-outcome combinations in the pair cohort yielded 134 associations that were significant after correction of false-discovery-rate (FDR) at *q* ≤ 0.05 (Fig. 1). Cesarean delivery was associated with higher diagnosis counts for the composite neurodevelopmental outcome (23.4%; 95% CI 21.1, 25.7), ASDs (20.0%; 11.5, 29.0), ADHD (10.4%; 6.5, 14.4), speech/language disorders (19.3%; 16.5, 22.1), developmental coordination disorder (21.7%; 15.1, 28.6), depression (9.1%; 4.7, 13.7), anxiety (3.8%; 0.9, 6.8), and elimination disorders (enuresis and encopresis; 7.6%; 3.4, 12.1). Maternal infection and immune-related diagnoses were also strong pair signals. Maternal immune-mediated disease was associated with higher composite neurodevelopmental (27.4%; 21.3, 33.7), ADHD (32.4%; 20.6, 45.3), and speech/language (24.0%; 16.7, 31.7) diagnosis counts.

**Figure 1:**
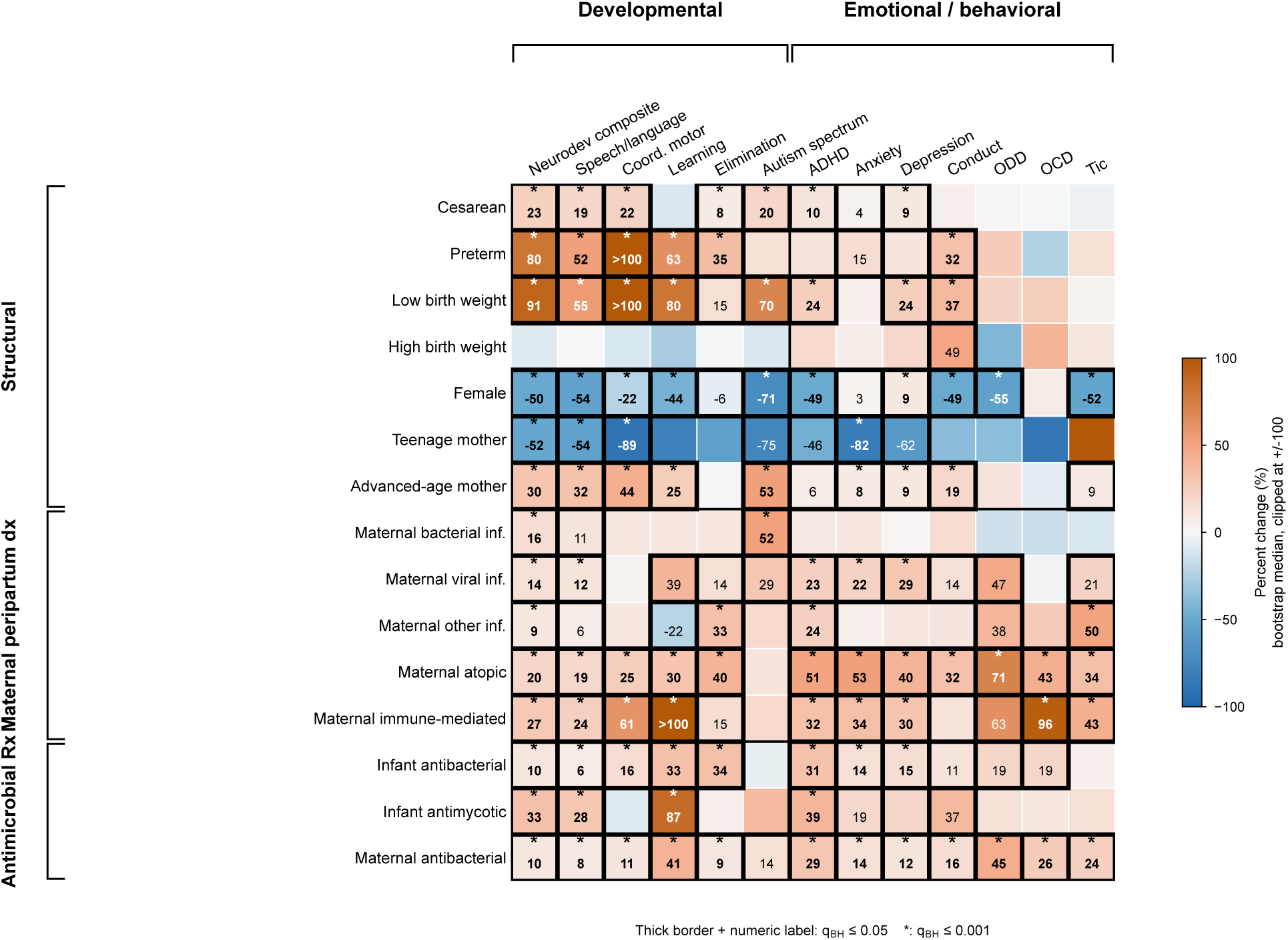
Population-level perinatal-exposure atlas across 13 DSM-5 psychiatric and neurodevelopmental outcomes. Percent change in expected childhood-diagnosis counts per unit exposure (negative-binomial regression adjusted for birth year with fitted-model coefficient co-variance propagated through 40,000 bootstrap draws; analytic cohort up to *n* = 1,179,611 mother-newborn pairs, with outcome-specific zero-event birth-year exclusions reducing *n* for sparse outcomes; intellectual disability excluded due to theta-boundary non-convergence). Rows show 15 displayed perinatal exposures grouped by domain; columns show 13 DSM-5 outcomes arranged by developmental versus emotional/behavioral classification. In figure labels, elimination disorders denote enuresis and encopresis. Outlined cells with numeric labels are significant after Benjamini-Hochberg FDR adjustment across the 234-cell primary matrix (*q* ≤ 0.05); asterisk denotes *q* ≤ 0.001. Color is clipped from −100% to 100% to preserve resolution near zero. The infant antiparasitic, maternal antimycotic, and maternal antiparasitic exposure rows are excluded from this main display because of small numerator counts and consequent confidence-interval instability that produced clipped color cells without inferential support; full estimates for these rows are reported in the population main-arm aggregate source table. Preterm birth and low birth weight show broad, generally larger population effects than cesarean; the cesarean effect is present on several developmental outcomes and ADHD but weaker or absent on rarer emotional-behavioral outcomes.

Maternal atopic disease was associated with a higher composite neurodevelopmental burden (20.1%; 16.4, 23.9) and ADHD (50.5%; 41.9, 59.7). Maternal bacterial and viral infections were associated with ASDs (52.0% and 29.2%, respectively) and with the composite neurodevelopmental outcome (16.2% and 14.2%). Advanced maternal age was another broad population signal, including composite neurodevelopmental (30.3%), autism spectrum (52.8%), speech/language (31.7%), and ADHD (6.0%). Preterm birth and low birth weight showed larger composite effects (79.9% and 91.2%), setting the scale for established fetal development risk.

### The sibling cohort corroborates the CD–neurodevelopmental disorders signal

We then analyzed the sibling cohort to test which signals persisted after accounting for stable maternal and family-level factors. The constructed non-twin sibling cohort contained 259,339 children in 123,926 families; after model-specific zero-event birth-year exclusions, the primary fitted cesarean support set contained 259,318 children in 123,917 families. In the sibling fixed-effects branch, 118 estimable exposure-outcome cells remained after non-identified cells were dropped; 37 were significant at FDR *q* ≤ 0.05 (Fig. 2; Supplementary Table S5).

**Figure 2:**
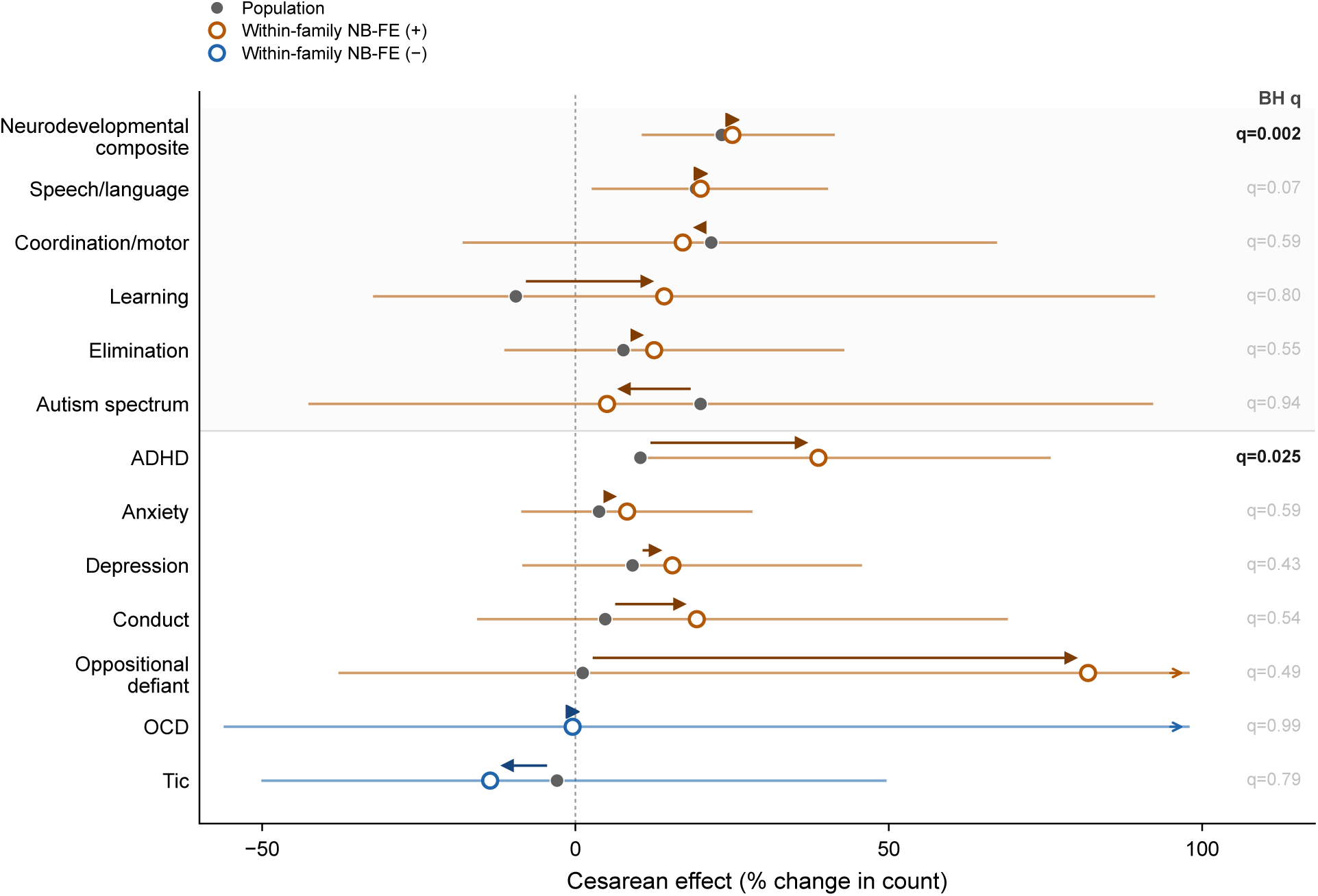
Within-family re-estimation across 13 outcomes: broad directional cesarean enrichment with concentrated individual-cell support. Population estimates (filled gray circles) and sibling negative-binomial fixed-effects estimates (open circles; orange for positive estimates, blue for negative estimates) for cesarean delivery across 13 outcomes (sibling fixed-effects construction cohort up to *n* = 259,339 non-twin children from 123,926 families; primary fitted cesarean support set *n* = 259,318 children in 123,917 families after model-specific zero-event birth-year exclusions). In figure labels, elimination disorders denote enuresis and encopresis. Horizontal intervals are 95% CIs for the sibling estimates; arrowheads mark CIs extending beyond the displayed axis. Cesarean point estimates are positive for 11 of 13 outcomes within families (descriptive exact-binomial sign-test reference *p* = 0.022; not a calibrated independent-outcome test). Individual NB-FE cells surviving BH-FDR at *q* ≤ 0.05: composite neurodevelopmental count (25.0%; *q* = 0.0015) and ADHD (38.8%; *q* = 0.025). Speech/language (20.0%; *q* = 0.070) is directional. Cesarean-on-autism is null (5.0%, *p* = 0.87).

CD remained a broadly predictive exposure in family-controlled analyses. As shown in Fig. 2 and Supplementary Table S5, the point estimates for the CD associations were positive for 11 of the 13 outcomes (descriptive sign-test reference *p* = 0.022). The strongest FDR support we observed was for CD–composite neurodevelopmental count (25.0%; 95% CI 10.6, 41.4; *q* = 0.0015) and ADHD (38.8%; 9.5, 75.8; *q* = 0.025). Speech/language disorder-related associations were positive but had weaker FDR support (20.0%; 2.6, 40.3; *q* = 0.070). The ASDs were the key boundary: the direction stayed positive, but the estimate within the family was small and non-significant (5.0%; 95% CI −42.6, 92.2; *p* = 0.87). Therefore, family-controlled results show a narrower signal than pair-cohort results: the population CD associations with composite neurodevelopmental burden and ADHD were reproduced within families, while the population CD–ASD association was not statistically supported in the smaller sibling design.

### Claims-defined no-labor/no-repeat cesarean carries the strongest developmental association

When CD was decomposed into priority-coded claims-defined subtypes, the no-labor/no-repeat category, the closest claims-data proxy for planned or pre-labor CD, showed larger coefficients than labor-onset CD for six developmental outcomes (Fig. 3): composite neurodevelopmental (ratio of rate ratios 1.24; 95% CI 1.19, 1.29), ASDs (1.31; 1.13, 1.51), specific learning disorders (1.63; 1.36, 1.95), speech/language (1.19; 1.14, 1.25), developmental coordination (1.30; 1.16, 1.45), and elimination disorders (1.26; 1.16, 1.36).

**Figure 3:**
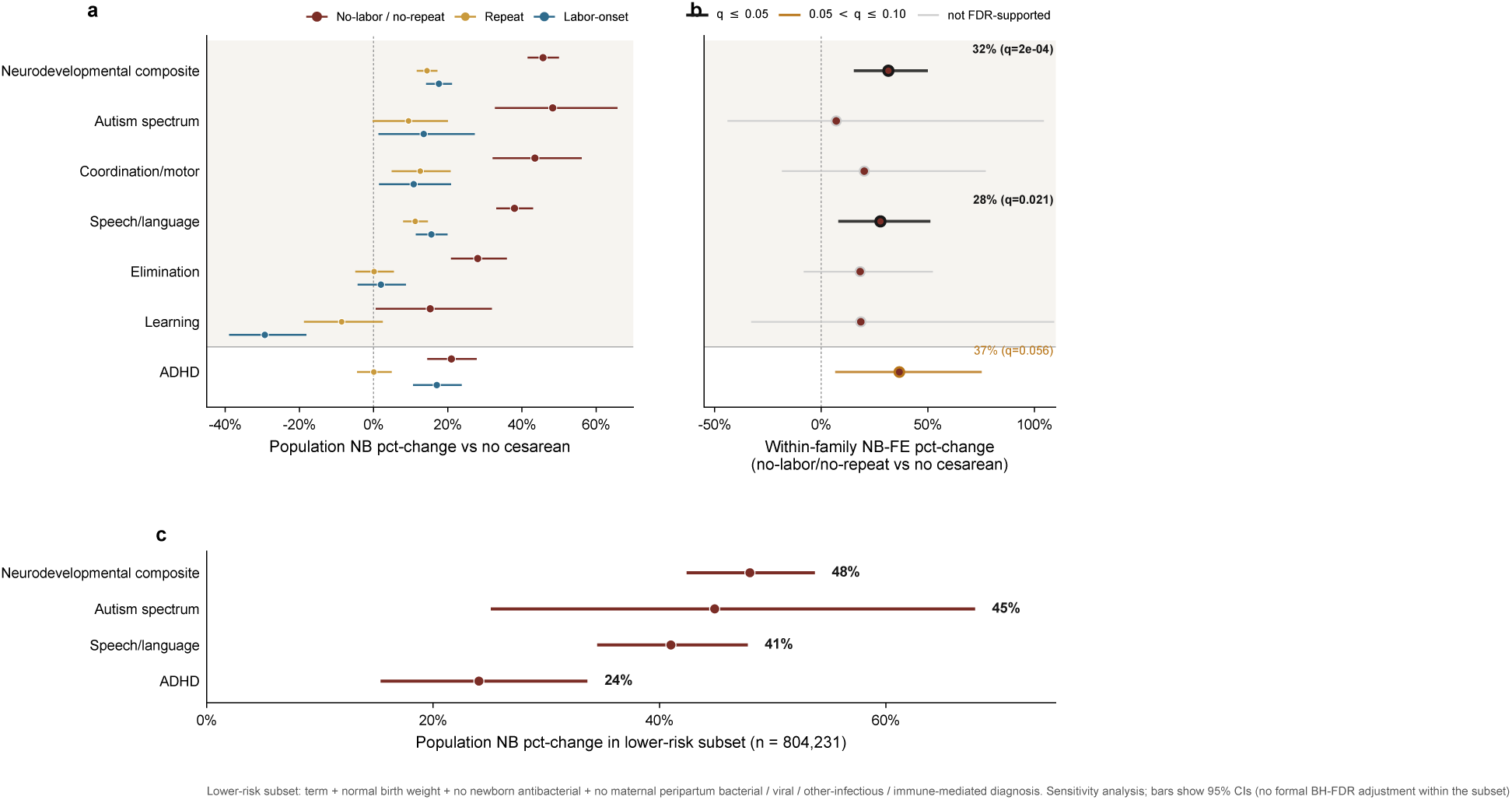
Cesarean subtype gradient: claims-defined no-labor/no-repeat cesarean carries the strongest developmental association. (a) Population negative-binomial percent change in expected diagnosis count, comparing claims-defined no-labor/no-repeat (red), repeat/prior-cesarean without detected labor-onset (tan), and claims-defined labor-onset (blue) cesarean to no cesarean, across six developmental outcomes and ADHD; horizontal intervals are 95% CIs from 40,000 MVN bootstrap draws on the decomposition-arm fits. In figure labels, elimination disorders denote enuresis and encopresis. The no-labor/no-repeat subtype has the largest estimate for each displayed developmental outcome; ADHD does not show the same subtype gradient. (b) Within-family negative-binomial mother fixed-effects percent change for the no-labor/no-repeat subtype on the same outcomes; points and 95% CIs are from sibling NB-FE fits. Black borders mark cells with BH-FDR *q* ≤ 0.05, amber borders mark 0.05 *< q* ≤ 0.10, gray borders mark unsupported cells. Within-family subtype support is concentrated in the composite neurodevelopmental count (31.5%; *q* = 0.00023) and speech/language disorders (27.8%; *q* = 0.021), with ADHD marginal (36.7%; *q* = 0.056). (c) Lower-risk population stress test for the no-labor/no-repeat subtype after restriction to term, normal-birth-weight children without newborn antibacterial dispensing and without maternal peripartum bacterial, viral, other-infectious, or immune-mediated diagnoses (*n* = 804,231). The claims-defined subtypes do not validate clinical intent; “no-labor/no-repeat” is the closest claims-data proxy for pre-labor or planned cesarean, and “labor-onset” the closest proxy for intrapartum cesarean.

Within families, claims-defined no-labor/no-repeat CD retained the strongest subtype association with composite neurodevelopmental count (31.5%; *q* = 0.00023) and speech/language disorders (27.8%; *q* = 0.021). The ADHD estimate was positive, but did not pass the primary FDR threshold (36.7%; *q* = 0.056). In a lower-risk population stress test that excluded preterm birth, low birth weight, antibacterial dispensing of the newborn, and maternal peripartum infection/immune markers (*n* = 804,231), claims-defined no-labor/no-repeat CD was associated with higher composite neurodevelopmental diagnoses (48.0%), ASDs (44.9%), speech/language (41.0%) and ADHD (24.1%). This analysis was a targeted sensitivity check, not an independent discovery analysis.

### Preterm birth and low birth weight predict larger risks

Our analysis of exposures to preterm birth or/and low birth weight reproduced the well-established findings ^24–27^. Each association was positive for all 13 outcomes within families (descriptive sign-test references *p* = 2.4 × 10^−4^). FDR-significant associations with preterm delivery included composite neurodevelopmental (92.6%), ASDs (143.6%), learning (125.6%), speech/language (89.8%), developmental coordination (256.8%), and elimination disorders (41.1%). FDR-significant associations for low-birth-weight exposure included composite neurodevelopmental count (164.0%), speech/language (113.7%), and developmental coordination (210.9%). Thus, CD, preterm birth, and low birth weight all showed broad positive within-family directionality, but prematurity and low birth weight had larger and more uniformly significant developmental effects; the CD profile was smaller and more concentrated in neurodevelopmental burden and ADHD (Fig. 4).

**Figure 4:**
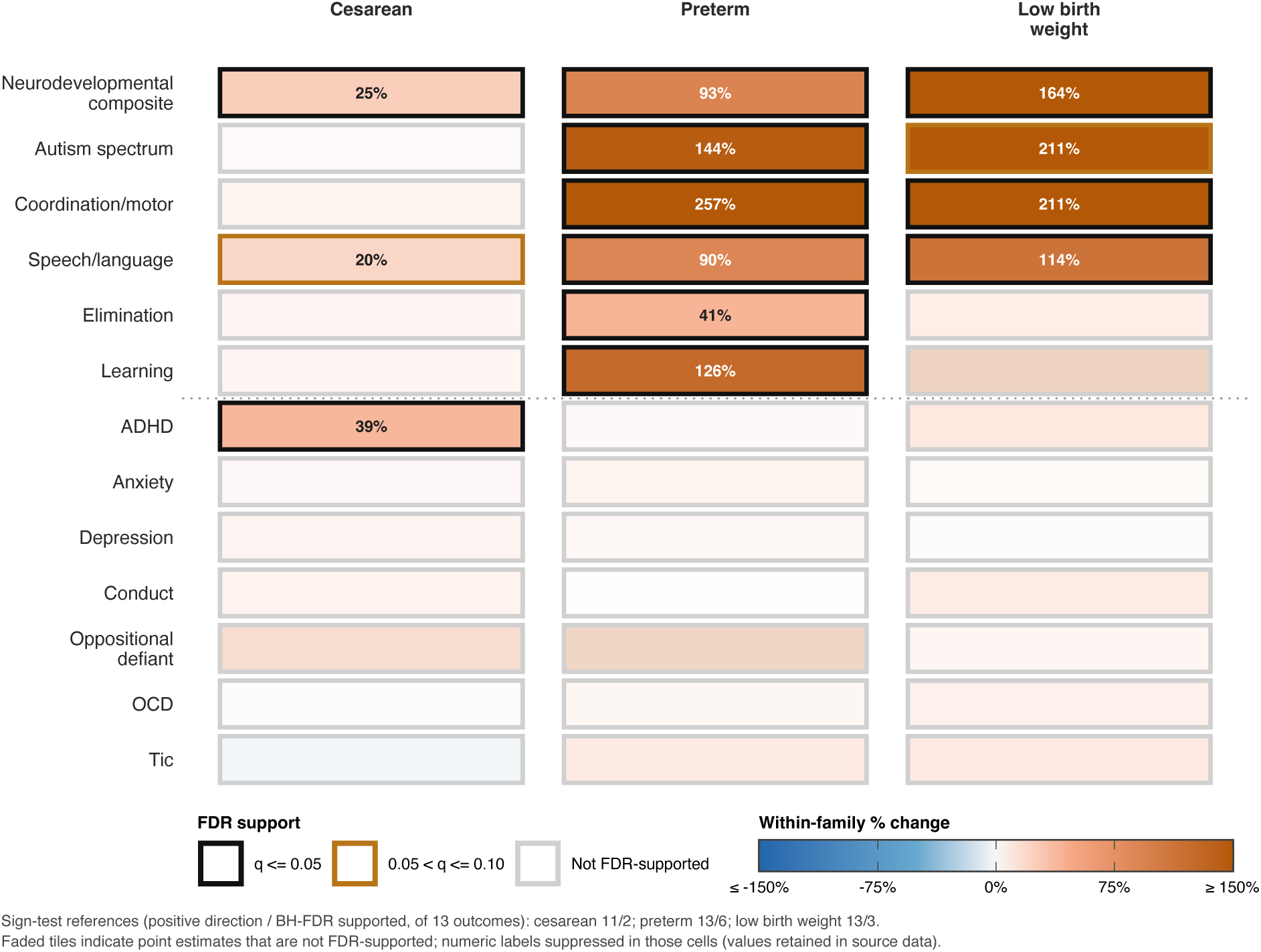
Within-family risk signatures separate the cesarean pattern from prematurity and fetal-growth risk. Tile plot of negative-binomial mother fixed-effects percent-change estimates for cesarean delivery, preterm birth, and low birth weight across the 13 DSM-5 outcome counts. In figure labels, elimination disorders denote enuresis and encopresis. Column headers give the descriptive sign-count reference across outcomes (cesarean 11/13 positive; preterm 13/13 positive; low birth weight 13/13 positive). Cell labels show percent change; asterisks mark BH-FDR *q* ≤ 0.05. Black borders mark BH-FDR-supported cells, amber borders mark 0.05 *< q* ≤ 0.10, and gray borders mark unsupported cells. Exact-binomial sign-test reference values are reported for orientation only (cesarean *p* = 0.022; preterm *p* = 2.4 × 10^−4^; low birth weight *p* = 2.4 × 10^−4^) and are not calibrated independent-outcome tests. Cesarean shows broad directionality but smaller and less uniformly significant effects; preterm birth and low birth weight show larger developmental effects and a more uniformly positive within-family profile.

### Sensitivity analyses define the boundary of the cesarean claim

The CD exposure associations remain stable in several targeted stress tests (Fig. 5). Adding six observed maternal psychiatric/neurodevelopmental claims-history indicators left CD–developmental disorders association coefficients nearly unchanged (composite count 23.4% → 22.1%; speech/language 19.3% → 18.3%; developmental coordination 21.7% → 20.8%), while ADHD attenuated more (10.4% → 5.9%). Adjusting for the intensity of pediatric visits to newborns changed the CD–outcome association coefficients by ≤ 1.5 percentage points.

**Figure 5:**
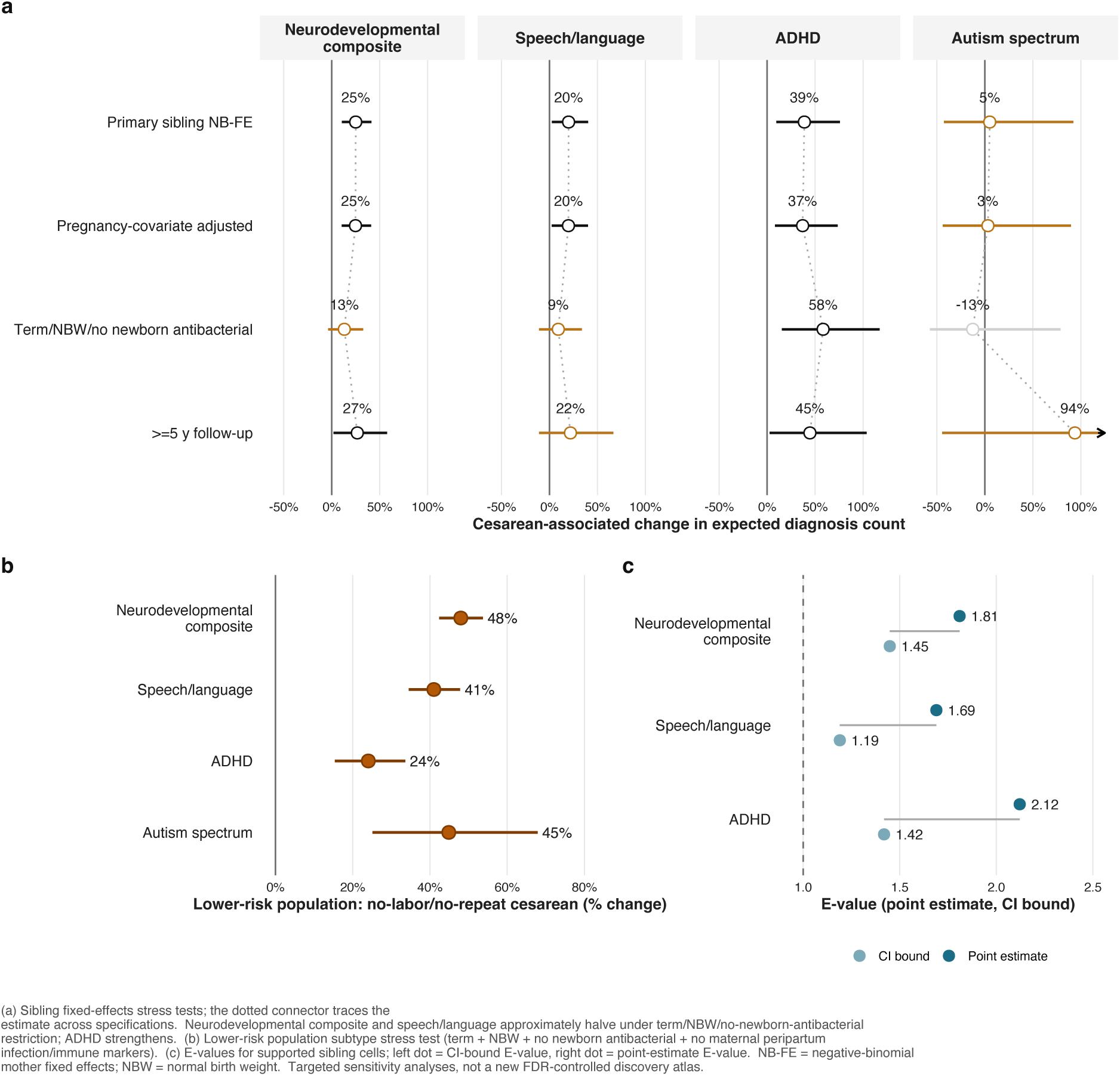
Cesarean robustness and causal-boundary analyses. (a) Sibling negative-binomial mother fixed-effects estimates for cesarean across four headline outcomes (composite neurodevelopmental, speech/language, ADHD, autism spectrum) under four specifications: primary, pregnancy-covariate adjusted, term + normal-birth-weight + no newborn antibacterial dispensing, and restriction to children with at least five years of follow-up. Dotted connectors trace each estimate across specifications. The composite neurodevelopmental and speech/language coefficients approximately halve under the term/normal-birth-weight/no-newborn-antibacterial restriction (composite 25% → 13%; speech/language 20% → 9%); the ADHD coefficient strengthens (39% → 58%; *p* = 0.0044 in the targeted restricted-cohort fit) and the autism coefficient remains imprecise. Arrowheads mark intervals extending beyond the displayed axis. (b) Lower-risk population subtype stress test restricted to term, normal-birth-weight children without newborn antibacterial dispensing and without maternal peripartum bacterial, viral, other-infectious, or immune-mediated diagnoses; estimates are for claims-defined no-labor/no-repeat cesarean. (c) E-values for the supported primary sibling cesarean cells; the left dot shows the CI-bound E-value and the right dot shows the point-estimate E-value. These are targeted sensitivity analyses, not a new FDR-controlled discovery atlas.

Restriction to full-term children of normal-birth-weight without exposure to antibacterial medications preserved the population CD estimates for composite neurodevelopmental count (23.8%), speech/language (20.8%), ADHD (11.2%) and autism spectrum (19.4%). The analogous sibling restriction was more heterogeneous: composite neurodevelopmental and speech/language estimates attenuated and lost nominal support (25.0% → 13.2%, *p* = 0.13; 20.0% → 9.2%, *p* = 0.39), ADHD strengthened (38.8% → 58.3%; 95% CI 15.4, 117.2; *p* = 0.0044), and autism remained imprecise (5.0% → −12.6%). Adding covariates of maternal-age, infectious, immune-mediated, and antimicrobial medications during pregnancy to the sibling models left the main cesarean pattern similar: composite 24.8%, speech/language 19.9%, ADHD 37.0%, and ASDs 3.2%. ICD-era stratification preserved the main developmental pattern but showed outcome-specific heterogeneity for learning, oppositional defiant disorder, OCD, and tic outcomes (Extended Data Fig. 2).

The value *E* is defined as the minimum strength of association that an unmeasured confounder would need to have with both exposure and outcome (on the risk-ratio scale), conditional on the measured covariates, to reduce the observed effect estimate to 1 (no association). The *E*-values for the CD associations were modest: composite neurodevelopmental *E* = 1.81 (CI-bound 1.45), ADHD *E* = 2.12 (CI-bound 1.42) and speech/language *E* = 1.69 (CI-bound 1.19). A negative-control probe within-family injury across high-preterm surveillance, low-birth weight, and newborn-antibacterial discordance defined an empirical ascertainment-bias floor of 1.18 to 1.30 (Extended Data Fig. 3).

### Neonatal antibacterial timing separates early developmental disorders from ADHD patterns

Neonatal antibacterial medication was not the main focus of our study, but provided a timing check for our analysis (Extended Data Fig. 5). Early-life antibiotic associations are vulnerable to indication, surveillance, and protopathic bias ^28–31^. The composite neurodevelopmental association was strongest in the first 28 days after birth and then declined (32.5% at 0–28 days, 24.7% at 29–90 days, 6.0% at 91–182 days; earliest-window factor linear component *p* = 5.3×10^−9^). ADHD was less interpretable: the mutually adjusted binary-window estimates were 14.1%, 23.2%, and 28.8%, but the separate earliest-window factor model was non-monotone (RRs 1.35, 1.41, 1.29; empirical peak 29–90 d), despite a strong linear component (*p* = 8.9×10^−32^) and significant deviation from linearity (*p* = 4.17 × 10^−4^). We therefore interpret the ADHD timing pattern as compatible with family-level confounding, surveillance, or reverse causation, not as a clean ascending exposure-window gradient.

## Discussion

This study identified a broad perinatal exposure signature of childhood neurodevelopmental and psychiatric diagnosis in a national commercial-claims cohort. Maternal immune-mediated disease, maternal infection, advanced maternal age, preterm birth, low birth weight, and cesarean delivery showed population-level associations. This was a biologically motivated atlas of plausible perinatal exposures, not a hypothesis-free search across all claims. The sibling analysis then sharpened the result: after mother fixed effects, cesarean delivery retained a broad directional pattern, with the strongest support for composite neurodevelopmental burden and ADHD, while ASDs were not statistically supported by analysis of the smaller sibling cohort.

The diagnosis-day outcomes were sparse for most children, but heavy-tailed among children with complex or high-intensity care, producing overdispersed count distributions. We therefore used negative-binomial count models with person-time offsets and outcome-specific dispersion parameters so that the variance was not constrained to equal the mean.

The two-cohort design in our analysis is important. The pairs cohort tells us what can be detected at population scale. The sibling cohort asks which signals survive the comparison among children born to the same mother. Under that stricter design, cesarean delivery remained associated with composite neurodevelopmental count (25.0%) and ADHD (38.8%), but not with autism spectrum disorders (Fig. 2; Supplementary Table S5). This pattern is more informative than a single ASDs or ADHD result: it separates a robust neurodevelopmental/ADHD signal from population associations that may reflect family structure, pregnancy indications, surveillance, or other factors that vary in time. Reproduction of the CD–composite neurodevelopmental and CD–ADHD signals across the population and sibling cohorts supports those two associations; the absence of statistically significant autism support in the smaller sibling cohort should not be interpreted as evidence that there is no weaker association of ASDs.

Advanced maternal age was strongly associated with autism spectrum diagnoses in the population model, whereas teenage maternal age was associated with lower autism-spectrum diagnosis counts. Because these estimates come from observational claims data and may reflect ascertainment, cohort, socioeconomic, and family-structure differences, they should not be interpreted as evidence that maternal-age distributions explain secular autism-prevalence trends.

The no-labor/no-repeat cesarean analysis helps localize the cesarean signal. The subtype pattern was strongest for developmental outcomes and weaker or absent for ADHD and emotional/behavioral diagnoses, consistent with a planned or pre-labor cesarean hypothesis but without demonstrating clinical intent. In the lower-risk population stress test, claims-defined no-labor/no-repeat CD estimates were 48.0% for composite neurodevelopmental burden, 44.9% for ASDs, 41.0% for speech/language, and 24.1% for ADHD.

Our clinical interpretation is tentative, but straightforward. Cesarean delivery is a common and often elective procedure ^32^, and even modest associations matter on a population scale. At the same time, preterm birth and low birth weight remain much stronger risk factors when analyzed in the sibling cohort, but these exposures are never elective. Cesarean delivery should be treated as a marker of developmental risk that warrants follow-up analyses and mechanistic investigation.

Maternal immune-mediated disease and maternal infection exposures were also prominent in the pair cohort, especially in terms of neurodevelopmental burden, ADHD, speech/language disorders, and ASDs. These findings fit a broader framework of developmental-immunology ^3,5^, but this study does not provide the same primary sibling-controlled evidence for those maternal exposures as it does for CD. Maternal infectious, immune-mediated, atopic, and antimicrobial variables entered the targeted CD sensitivity models as pregnancy-varying covariates rather than as a full sibling-discovery matrix. They remain important population signals and strong candidates for future sibling-discordant, within-pregnancy, or biomarker-linked analyses.

The ASDs results are a useful boundary between results supported by both cohort analyses and unsupported associations. The pair cohort suggested a positive association of CD-ASDs, but the estimate derived from the smaller-size sibling cohort was small and, although consistent in direction with the pair cohort analysis, did not differ significantly from zero. That resonates with the caution provided by previous studies on population estimates, sibling comparison, and family risk studies ^14,33^. The cross-cohort comparison is shown in Extended Data Fig. 4. The outcome analysis for ADHD is different: our estimate using the sibling cohort was positive but did not show the same risk difference between the subtype of CD with no-labor/no-repeat, and it is higher than Curran et al.’s elective and emergency sibling-matched estimates ^15^. We therefore treat the

ADHD result as a cohort-specific signal that requires replication rather than as settled concordance with prior family-comparison evidence. Previous work on ADHD has also emphasized the need for genetically informed or family-comparison designs^15,31^.

Putting all our observations together, we arrive at a hypothetical mechanistic explanation. Our results suggest that neonatal exposures associated with changes in the maternal and newborn’s microbiomes (medications, infections, mode of delivery) affect neurodevelopmental and psychiatric traits of the child. The gut–brain axis operates from early life^34^: in preterm infants, dysbiosis associated with necrotizing enterocolitis, sepsis, and heavy antibiotic exposure correlates with worse cognitive and motor outcomes ^35,36^, likely mediated by systemic inflammation and altered microbial metabolites^37^. Mechanistically, communication proceeds along several parallel routes: direct neural signaling through the vagus nerve, whose afferent fibers sense bacterial products and metabolites via enteroendocrine cells and neuropods^38,39^; humoral transport of microbially produced short-chain fatty acids (SCFAs)—principally acetate, propionate, and butyrate—which cross the blood–brain barrier, regulate tight-junction integrity, and govern microglial maturation and homeostasis^40–42^; and modulation of the tryptophan–serotonin and kynurenine pathways, through which gut microbes shape neurotransmitter precursor availability and neuroimmune tone ^43,44^. These same metabolite classes also regulate the hypothalamic–pituitary–adrenal axis and microglial inflammatory programs during the perinatal window, when both the gut microbiome and the brain undergo their most rapid co-development^45,46^. Early microbiota disturbances are associated with stunted growth and undernutrition in low-resource settings, suggesting a role in energy harvest, nutrient absorption, and growth-factor signaling. Our results suggest the exciting possibility that an early neonatal probiotic intervention might be able to reduce the risk of neurodevelopmental and psychiatric childhood disorders. As probiotics can be designed to be minimally invasive, one can imagine their routine administration, which can potentially reduce the total burden of childhood-onset psychiatric and neurodevelopmental disorders. The hypothesis is experimentally and clinically testable.

Our study has several limitations. MarketScan includes records for more than two thirds of the US population over time, but the analytic cohort in this study represents commercially insured US individuals and excludes Medicaid beneficiaries and the uninsured. Follow-up after the exposure window was narrow (mean 3.43 years; median 2.35), allowing only early-onset diagnoses as outcomes. Administrative data also encode what is observed in care, not necessarily the underlying disease ^47,48^. The mother fixed-effects analysis removes stable family-level confounders but not carry-over effects, exposure measurement error, or changes in delivery planning or clinical indication^10–12^. The CD exposure estimates derived from the sibling cohort were calculated only from discordant families (10.6% of the sibling families), so the statistical power for detecting signals was further limited. Descriptive sign-test references were not calibrated with independent-outcome tests. The CD subtype decomposition depended on the accuracy of the insurance claims coding; *E*-values were modest in magnitude.

The next studies, data permitting, should move from association to causal identification: obstetric instruments such as breech presentation for planned CD^49^, term and preterm-stratified sibling analyses, richer maternal infection and immune phenotyping, and linkage to maternal body mass index, fetal growth, microbiome, umbilical cord-blood immune markers or newborn inflammatory profiles ^3,5,6,8^. The present study defines the target: maternal immune and delivery-mode signals are visible on the population scale, and cesarean delivery shows the clearest family-controlled association with childhood neurodevelopmental burden and ADHD.

## Online Methods

### Cohort construction

We identified mother-newborn pairs in Merative MarketScan Commercial Claims^50^ covering 2003 to 2024 by linking newborn enrollees to their maternal enrollee record through the MarketScan family linkage field. We required at least 365 days of maternal enrollment in the pre-birth interval and at least 365 days of newborn enrollment beginning at birth, with a 31-day tolerance for single-day coverage gaps. Births were defined by the presence of any inpatient or outpatient labor-delivery claim in the maternal record with a matched newborn enrollment within 31 days of the delivery claim. The resulting cohort comprised 1,179,611 mother-newborn pairs with 4,041,343 person-years of follow-up (Extended Data Fig. 1).

### Exposures

The 18 primary perinatal exposures are binary or categorical. Cesarean delivery was defined from claims-based maternal delivery diagnoses near birth, newborn delivery-type diagnoses, and procedure-code evidence (CPT plus ICD-9 procedure-code prefixes) within 30 days after delivery. A secondary cesarean decomposition assigned each cesarean delivery to one of three mutually exclusive priority-coded claims-defined subtypes: no-labor/no-repeat cesarean (no detected delivery-window labor-onset code and no delivery-window repeat/prior-cesarean diagnosis code), repeat/prior-cesarean without detected labor-onset (delivery-window repeat/prior-cesarean diagnosis code with no detected labor-onset code after priority remapping), or labor-onset cesarean (detected labor-onset code). Because labor-onset and repeat/prior-cesarean evidence can co-occur in claims, the priority was labor-onset *>* repeat/prior-cesarean *>* no labor/no repeat. These are categories defined by claims rather than validated planned-delivery or clinical-urgency phenotypes. Preterm birth was defined by an ICD-coded gestational-age diagnosis before 37 weeks. The birth-weight categories (low *<* 2500 g; high ≥ 4500 g) were defined by ICD-coded birth-weight diagnosis. Maternal age indicators were defined from birth-year-derived maternal age as teenage mother (13–18 years) and advanced-age mother (35–45 years). Maternal peripartum exposures (bacterial, viral, other-infectious, atopic, immune-mediated diagnosis categories) were defined by any ICD codes in the matching category in the maternal record within 270 days before the newborn’s birth. Neonatal and maternal antimicrobial dispensing (antibacterial, antimycotic, antiparasitic) was defined as the presence of at least one outpatient filled prescription mapped from NDC to the corresponding anti-infectious therapeutic class AHFS within the indicated window. For the maternal-psych arm, six additional indicators (maternal autism, ADHD, depression, anxiety, bipolar disorder, and psychotic disorders) were extracted as ever-observed maternal claims-history flags across MarketScan enrollment. For the surveillance arm, a three-level factor of newborn diagnostic-contact days was added in the first 183 days of life (1, 2–3 or 4+ days versus 0).

### Outcomes

The final outcome panel contained 13 DSM-5 psychiatric and neurodevelopmental outcome counts: composite neurodevelopmental-disorder count, autism spectrum disorder ^51,52^, ADHD ^53^, conduct disorder, oppositional defiant disorder, anxiety disorders, obsessive-compulsive disorder, tic disorders, specific learning disorders, speech/language disorders, developmental coordination disorder, depressive disorders, and elimination disorders (enuresis and encopresis). Each outcome is the count of distinct diagnosis dates in the respective ICD-9 / ICD-10 category appearing on the child’s claim record beginning 183 days after birth. Intellectual disability was defined in the outcome panel but excluded from population negative-binomial fits because MASS::glm.nb failed to converge at the theta boundary (641 children with at least one diagnosis; 4,880 diagnosis-days in 1.18 M rows); the result was retained in the within-family branch where fixest::fenegbin successfully converged.

### Population negative-binomial models (4 arms)

The main arm fit a negative binomial regression of each outcome count on the 18 exposures, a birth-year factor for the secular trend, and an offset of log(person_time). This specification was chosen for over-dispersed claims-count outcomes because the outcome-specific dispersion parameter allows the count variance to exceed the mean. For child *i* and outcome *k*, the population model was

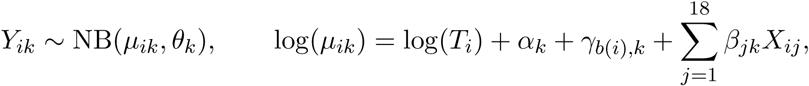

where *T_i_* is the observed person-time after the 183-day outcome washout, *θ_k_* is the outcome-specific dispersion parameter, *γ_b_*_(_*_i_*_)_*_,k_* is a birth-year effect and *X_ij_* are indicators of perinatal exposure. The model was fit via MASS::glm.nb^54^. Inference used a multivariate-normal parametric bootstrap of the coefficient vector with 40,000 draws, seed 42, using the fitted-model coefficient covariance in the aggregate source-data files. Ninety-five percent confidence intervals are empirical quantiles {2.5, 97.5} of the bootstrapped percent changes, 100{exp(*β_jk_*) − 1}. These population-branch intervals remain conventional model-based cohort estimates rather than within-family estimates; the sibling branch is the primary family-controlled inferential arm for exposures that vary between siblings. The surveillance arm added the three-level factor of newborn-diagnostic-contact; the decomposition arm replaced the single cesarean delivery indicator with the three subtype indicators; the maternal psychiatric arm added the six indicators of the history of maternal psychiatric/neurodevelopmental claims. All four arms were otherwise identical.

### Sibling fixed-effects models

Mothers with two or more births in the cohort were identified using the MarketScan family identifier. Twin pairs (siblings with identical birth days and a shared mother identifier) were excluded. The resulting within-family construction cohort comprised 259,339 children in 123,926 families. Outcome-specific fixed-effect fits then dropped birth-year strata with zero events; the primary cesarean fitted support set reported in sensitivity tables contained 259,318 children in 123,917 families. Negative-binomial fixed effects were fit by fixest::fenegbin^55^. For child *i* in mother-family *m* and outcome *k*, the sibling model was

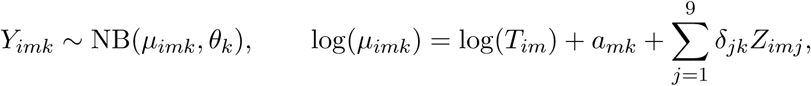

where *θ_k_* is the outcome-specific dispersion parameter, *a_mk_* is the absorbed mother fixed effect, and *Z_imj_* are time-varying child or pregnancy exposures: cesarean delivery, preterm birth, low birth weight, high birth weight, infant sex, birth order, and the three neonatal antimicrobial dispensing indicators. Under the decomposition arm, the single cesarean indicator was replaced by the three mutually exclusive subtypes. Standard errors were clustered by mother via fixest’s cluster-robust estimator ^56^.

### Descriptive sign-test references across the 13-outcome panel

For each exposure (cesarean, preterm, low birth weight), we counted positive point estimates in the 13 DSM-5 outcomes and computed the reference values of the exact-binomial sign-test under *H*_0_ : *p* = 0.5 via scipy.stats.binomtest. These values are reported only as descriptive orientation because the outcomes are correlated and the composite neurodevelopmental count overlaps component outcomes; they are not treated as calibrated independent-outcome tests or as replacements for individual-cell BH-FDR correction.

### Multiple testing

The empirical two-sided *p*-values were computed from the bootstrap drawings as *p* = 2×min(Pr(*β*^^^ *<* 0), Pr(*β*^^^ *>* 0)) + Pr(*β*^^^ = 0), bounded below at 1*/*(2*n*) to reflect the Monte Carlo resolution of the *n* = 40,000 drawings. Benjamini-Hochberg FDR correction^57^ was applied per arm in the primary matrix of each arm: 234 cells (18 exposures × 13 results) for the main arm, 273 for surveillance (adds the newborn-diagnostic-contact factor), 260 for decomposition (three cesarean subtypes replacing the single indicator), 312 for maternal-psych (adds six indicators of psychiatric-history). For the within-family branch, BH-FDR was applied in 118 cells (NB-FE main; nine exposures in the 13 displayed results plus intellectual disability where the model converged, less non-identified cells) and 146 (NB-FE decomposition; 11 exposures including three cesarean subtypes in the 13 displayed outcomes plus intellectual disability where converged, less non-identified cells).

### Cesarean sensitivity analyses

We ran targeted cesarean sensitivity analyses after the main atlas. First, population models were refitted after restricting to term births, term plus normal-birth-weight births, term plus normal-birth-weight births without newborn antibacterial dispensing, and children with at least two, three, or five years of post-birth observation. Second, the fixed effect models of the siblings were refitted under the same restrictions for the six main cesarean outcomes (composite neurodevelopmental, speech/language, ADHD, autism spectrum, developmental coordination and elimination disorders). Third, the main sibling cesarean models were re-fitted with additional pregnancy-varying covariates: maternal-age indicators (teenage mother, advanced-age mother), maternal peripartum bacterial, viral, other-infectious, atopic and immune-mediated diagnosis indicators, and maternal antimicrobial dispensing indicators. Fourth, a lower-risk cesarean-decomposition model restricted the population cohort to term children of normal-birth-weight without antibacterial medications exposure to newborns and without bacterial, viral, other-infectious or immune-mediated maternal peripartum diagnoses. These sensitivity analyses were not FDR-controlled as a new discovery atlas; they were targeted robustness analyses of headline cesarean claims.

### E-values

The *E*-values were computed per VanderWeele and Ding^58^: for a rate ratio greater than 1, 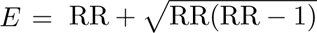; the CI-based *E*-value is from the 95% CI bound closest to the null. The key E-values within the family are reported in the Supplementary Table S7.

### Timing-window analysis of neonatal antibacterial dispensing

For neonatal antibacterial dispensing, whose timing is resolvable in the claims record from the fill date, we refit every population model with three mutually adjusted binary window indicators (0 to 28, 29 to 90, 91 to 182 days) replacing the single composite indicator and included the newborn diagnostic-contact factor when available. These indicators are not mutually exclusive: children exposed to medications in multiple windows can contribute to multiple window coefficients. A separate earliest-window factor (none, 0 to 28 days, 29 to 90 days, or 91 to 182 days) supported nested likelihood-ratio tests for an omnibus (3 df), linear-trend (1 df), and deviation-from-linearity (2 df) component.

### Code, data, reproducibility

All analysis code has been assembled in a private GitHub repository for reviewer access and will be made public on acceptance. Releasable aggregate outputs used for figures are supplied as Nature source-data workbooks with the submission and will be released with the repository on acceptance. Seeded bootstraps use seed 42. The code enforces cesarean-decomposition priority (labor-onset *>* repeat/prior-cesarean *>* no labor/no repeat) and row-level consistency checks. The Merative MarketScan individual-level data are gated by institutional license and cannot be redistributed. The exact NDC-to-AHFS anti-infective mapping file used to construct maternal and newborn antimicrobial indicators is not redistributed; its expected schema and reconstruction requirements from licensed NDC/AHFS resources are documented in the reviewer-access repository.

## Author contributions

Contributions are reported according to the CRediT taxonomy. **B.K.**: conceptualization, methodology, software, formal analysis, data curation, investigation, validation, visualization, writing – original draft, writing – review & editing. **S.A.K.**: supervision, resources, writing – review & editing. **A.R.**: conceptualization (originating study concept), supervision, resources, writing – review & editing. All authors approved the final manuscript.

## Data availability

Merative MarketScan Commercial Claims data are licensed from Merative and cannot be redistributed by the authors. Individual-level data access requires institutional licensing with Merative. Derived summary statistics that do not contain individual-level claims records (figure source data, result-table summaries, and aggregated cohort descriptives) are supplied with the submission where licensing permits and will be released with the public repository on acceptance. Raw reconstruction of the antimicrobial exposure branch also requires access to the licensed NDC/AHFS mapping resource documented in the reviewer-access repository.

## Code availability

All analysis code (cohort construction, exposure and outcome definitions, individual-level and sibling fixed-effects models, timing-window analysis, figure generation) has been assembled in a private GitHub repository for reviewer access and will be released publicly on acceptance. The analytic environment is captured in the repository, including R 4.5.3 with the fixest, MASS, and sandwich packages and Python 3.11 dependencies.

## Ethics statement

This study used fully de-identified Merative MarketScan Commercial Claims data under the University of Chicago’s HIPAA-compliant data-use agreement with Merative. The University of Chicago Biological Sciences Division Institutional Review Board determined the study exempt from IRB review under the secondary-research use of de-identified data exemption (45 CFR 46.104(d)(4)).

## Supporting information

Complete Supplementals

## Acknowledgments

We thank the Merative MarketScan team for data access.

