## Supplementary material for "Maternal immunity, cesarean delivery, and childhood neuropsychiatric risk in 1.18 million births": Complete Supplementals

10 May 2026

#### Contents

**1 Analysis Arms and FDR Denominators** 2

**2 Cohort and Cesarean Subtype Counts** 3

**3 Population-Level Headline Effects** 4

**4 Within-Family Headline Effects** 5

**5 Within-Family Cesarean Sensitivity Grid** 6

**6 Cesarean Decomposition** 8

**7 E-Values** 9

**8 Negative-Binomial Dispersion Diagnostics** 10

**9 Cesarean Sensitivity Analyses** 11

**10 Timing and Differential-Surveillance Checks** 13

**11 Data and Code Availability** 14

### 1 Analysis Arms and FDR Denominators

Table S1: **Analytic arms and Benjamini–Hochberg FDR counts.** Significant counts use the arm-specific primary BH-FDR threshold of $q \leq 0.05$. Denominators correspond to the model families used for FDR correction.

| Arm | Primary matrix | Significant | Total |
|---|---|---|---|
| Population main | 18 exposures × 13 outcomes | 134 | 234 |
| Population surveillance-adjusted | Main arm plus newborn diagnostic-contact factor | 160 | 273 |
| Population cesarean decomposition | Three populated claims-defined cesarean subtype indicators replacing the single cesarean indicator | 154 | 260 |
| Population maternal-psych | Main arm plus six maternal psychiatric-history indicators | 169 | 312 |
| Sibling NB-FE | Nine time-varying exposures across the 13 displayed outcomes plus intellectual disability where converged, less non-identified cells | 37 | 118 |
| Sibling NB-FE cesarean decomposition | Eleven time-varying exposures including three cesarean subtypes across the 13 displayed outcomes plus intellectual disability where converged | 33 | 146 |

#### 2 Cohort and Cesarean Subtype Counts

Table S2: **Cohort and claims-defined cesarean subtype counts.** Percentages are column percentages within the full analytic cohort ($n$ = 1,179,611) or the final non-twin sibling FE construction cohort ($n$ = 259,339). The priority remap assigns every cesarean delivery to no-labor/no-repeat, repeat/prior-cesarean without detected labor-onset, or labor-onset categories; the residual unspecified subtype is zero by construction after remapping and is not modeled.

| | **Full cohort** | | **Sibling FE cohort** | |
|---|---|---|---|---|
| **Measure** | **n** | % | **n** | % |
| Children | 1,179,611 | 100.0 | 259,339 | 100.0 |
| Families | – | – | 123,926 | – |
| Person-years | 4,041,343 | – | 1,219,429 | – |
| Cesarean delivery, any | 433,171 | 36.7 | 81,705 | 31.5 |
| Claims-defined no-labor/no-repeat cesarean | 125,665 | 10.7 | 20,346 | 7.8 |
| Repeat/prior-cesarean without detected labor-onset | 191,595 | 16.2 | 40,778 | 15.7 |
| Claims-defined labor-onset cesarean | 115,911 | 9.8 | 20,581 | 7.9 |
| Residual unspecified cesarean | 0 | 0.0 | 0 | 0.0 |
| Preterm birth | 84,291 | 7.1 | 13,866 | 5.3 |
| Low birth weight | 46,978 | 4.0 | 6,755 | 2.6 |
| Newborn antibacterial dispensing | 170,428 | 14.4 | 38,832 | 15.0 |
| Newborn antimycotic dispensing | 15,614 | 1.3 | 3,448 | 1.3 |
| Newborn antiparasitic dispensing | 195 | 0.017 | 30 | 0.012 |

The sibling-construction workflow identified 277,324 candidate children. Excluding twins removed 16,986 rows; requiring at least two remaining non-twin children per mother removed 999 singleton remnants, leaving 259,339 children in 123,926 families. Cesarean-discordant families numbered 13,189, or 10.6% of the sibling FE families.

### 3 Population-Level Headline Effects

Table S3: **Population main-arm summary for cesarean, preterm birth, and low birth weight.** Positive and BH-significant counts are across the 13 modeled DSM-5 outcomes.

| Exposure | Positive | BH-significant | Selected estimates |
|---|---|---|---|
| Cesarean delivery | 10/13 | 8/13 | Composite neurodevelopmental 23.4% (95% CI 21.1, 25.7); autism 20.0% (11.5, 29.0); ADHD 10.4% (6.5, 14.4); learning −9.5% (−17.4, −0.9), $q = 0.055$. |
| Preterm birth | 12/13 | 7/13 | Composite neurodevelopmental 79.9% (70.4, 90.0); developmental coordination 172.7% (134.2, 217.6); OCD is the only negative point estimate (−23.8%), not BH-significant. |
| Low birth weight | 13/13 | 9/13 | Composite neurodevelopmental 91.2% (81.0, 101.9); autism 70.3% (37.7, 110.0); developmental coordination 133.7% (100.6, 172.6). |

Table S4: **Selected population main-arm maternal infection, immune, and maternal-age effects.** Values are percent changes in expected diagnosis counts from the primary population negative-binomial models.

| Exposure | Selected estimates |
|---|---|
| Maternal immune-mediated disease | Composite neurodevelopmental 27.4% (95% CI 21.3, 33.7); ADHD 32.4% (20.6, 45.3); speech/language 24.0% (16.7, 31.7). |
| Maternal atopic disease | Composite neurodevelopmental 20.1% (16.4, 23.9); ADHD 50.5% (41.9, 59.7). |
| Maternal bacterial infection | Autism spectrum 52.0% (22.5, 88.8); composite neurodevelopmental 16.2% (9.8, 22.9). |
| Maternal viral infection | Autism spectrum 29.2% (10.3, 50.7); composite neurodevelopmental 14.2% (9.7, 18.9). |
| Advanced maternal age | Composite neurodevelopmental 30.3% (27.9, 32.8); autism spectrum 52.8% (41.9, 64.3); speech/language 31.7% (28.6, 34.8); ADHD 6.0% (2.2, 9.9). |

#### 4 Within-Family Headline Effects

Table S5: **Within-family negative-binomial fixed-effects headline cells.** Percent changes and Wald 95% confidence intervals come from the per-outcome sibling fixed-effects model summaries. $q$ values are Benjamini–Hochberg adjusted over the 118-cell sibling NB-FE primary matrix.

| **Exposure** | **Outcome** | **% change** | **95% CI** | **BH $q$** |
|---|---|---|---|---|
| Cesarean | Composite neurodevelopmental | 25.0 | 10.6, 41.4 | 0.0015 |
| Cesarean | ADHD | 38.8 | 9.5, 75.8 | 0.025 |
| Cesarean | Speech/language | 20.0 | 2.6, 40.3 | 0.070 |
| Cesarean | Autism spectrum | 5.0 | −42.6, 92.2 | 0.936 |
| Preterm | Composite neurodevelopmental | 92.6 | 67.1, 121.8 | $8.9 \times 10^{-19}$ |
| Preterm | Autism spectrum | 143.6 | 25.4, 373.4 | 0.031 |
| Preterm | Specific learning | 125.6 | 27.4, 299.7 | 0.021 |
| Preterm | Speech/language | 89.8 | 57.9, 128.1 | $5.9 \times 10^{-11}$ |
| Preterm | Developmental coordination | 256.8 | 143.6, 422.7 | $3.9 \times 10^{-10}$ |
| Preterm | Elimination | 41.1 | 8.3, 83.7 | 0.036 |
| Low birth weight | Composite neurodevelopmental | 164.0 | 118.4, 219.0 | $8.5 \times 10^{-23}$ |
| Low birth weight | Speech/language | 113.7 | 67.7, 172.3 | $4.5 \times 10^{-9}$ |
| Low birth weight | Developmental coordination | 210.9 | 91.3, 405.3 | $2.1 \times 10^{-5}$ |
| Low birth weight | Autism spectrum | 210.9 | 15.3, 738.5 | 0.076 |
| Low birth weight | Specific learning | 118.1 | 3.1, 361.4 | 0.116 |
| Low birth weight | ADHD | 40.9 | −1.8, 102.2 | 0.154 |

Panel-level descriptive sign-test references across the 13-outcome panel: cesarean was positive for 11 of 13 outcomes under NB-FE (descriptive exact-binomial sign-test reference $p = 0.022$); preterm birth and low birth weight were each positive for all 13 outcomes (descriptive exact-binomial sign-test reference $p = 2.4 \times 10^{-4}$ for each). Because the outcomes are correlated and the composite neurodevelopmental count overlaps component outcomes, these reference values are descriptive rather than calibrated independent-outcome tests.

### 5 Within-Family Cesarean Sensitivity Grid

Table S6a: **Within-family cesarean sensitivity grid for the four headline outcomes.** Negative-binomial mother fixed-effects percent changes with 95% confidence intervals across four within-family specifications: primary, pregnancy-covariate adjusted, term plus normal birth weight plus no newborn antibacterial dispensing, and at least five years of post-birth follow-up. The strengthening of the within-family ADHD coefficient under the term/normal-birth-weight/no-newborn-antibacterial restriction is the targeted-restriction finding referenced in the Results sensitivity paragraph. This targeted grid summarizes prespecified robustness checks and is not a separate discovery matrix.

| **Outcome** | **Primary** | **Preg-cov adj** | **Term + NBW + no abx** | **≥5 y follow-up** |
|---|---|---|---|---|
| Composite NDD | 25.0 [10.6, 41.4]* | 24.8 [10.4, 41.1]* | 13.2 [−3.6, 32.9] | 26.7 [1.8, 57.7]* |
| Speech / language | 20.0 [2.6, 40.3]* | 19.9 [2.5, 40.2]* | 9.2 [−10.8, 33.9] | 21.9 [−10.8, 66.7] |
| ADHD | 38.8 [9.5, 75.8]* | 37.0 [8.1, 73.5]* | **58.3** [15.4, 117.2]** | 44.6 [2.6, 103.7]* |
| Autism spectrum | 5.0 [−42.6, 92.2] | 3.2 [−43.9, 89.7] | −12.6 [−57.3, 78.9] | 93.8 [−44.5, 576.8] |
| $n$ | 259,318 | 259,318 | 175,462 | 59,023 |
| Families | 123,917 | 123,917 | 84,315 | 28,319 |
| Discordant families | 13,189 | 13,188 | 8,086 | 2,743 |

*Nominal $p < 0.05$. **$p = 0.0044$ in the targeted restricted-cohort fit. The primary fitted support set starts from the constructed non-twin sibling cohort of 259,339 children in 123,926 families; model-specific zero-event birth-year exclusions remove 21 children and 9 family identifiers from the displayed primary cesarean fits, leaving 259,318 children in 123,917 families. The pregnancy-covariate-adjusted column reports one fewer cesarean-discordant family than the primary column (13,188 vs 13,189) under the expanded covariate set.

### 6 Cesarean Decomposition

Table S6b: **Population no-labor/no-repeat-versus-labor-onset cesarean contrasts.** Values are ratios of rate ratios comparing claims-defined no-labor/no-repeat cesarean with claims-defined labor-onset cesarean, with 95% intervals and bootstrap probability that the no-labor/no-repeat coefficient exceeds the labor-onset coefficient.

| **Outcome** | **Ratio of RRs** | **95% CI** | **Pr** |
|---|---:|---:|---:|
| Composite neurodevelopmental | 1.239 | 1.192, 1.287 | 1.0000 |
| Autism spectrum | 1.306 | 1.128, 1.514 | 0.9999 |
| Specific learning | 1.631 | 1.358, 1.955 | 1.0000 |
| Speech/language | 1.194 | 1.139, 1.251 | 1.0000 |
| Developmental coordination | 1.295 | 1.161, 1.449 | 1.0000 |
| Elimination | 1.256 | 1.160, 1.360 | 1.0000 |
| Anxiety | 1.049 | 0.992, 1.110 | 0.9545 |
| ADHD | 1.034 | 0.962, 1.112 | 0.8172 |
| Conduct | 1.047 | 0.941, 1.164 | 0.7965 |
| Depression | 1.054 | 0.972, 1.144 | 0.8992 |
| OCD | 1.009 | 0.753, 1.358 | 0.5244 |
| Tic | 1.075 | 0.935, 1.232 | 0.8477 |
| ODD | 0.883 | 0.698, 1.111 | 0.1459 |

Within families, claims-defined no-labor/no-repeat cesarean was BH-significant for the composite neurodevelopmental count (31.5%, $q = 0.00023$) and speech/language disorders (27.8%, $q = 0.021$). The no-labor/no-repeat ADHD cell was positive but marginal (36.7%, $q = 0.056$). Repeat/prior-cesarean without detected labor-onset and labor-onset subtype rows had zero BH-significant within-family cells.

### 7 E-Values

Table S7: **E-values for key within-family cells.** E-values are computed from the sibling NB-FE rate ratio and the 95% confidence-bound nearest the null.

| **Exposure** | **Outcome** | **RR** | **E-value** | **CI-bound E-value** |
|---|---|---|---|---|
| Cesarean | Composite neurodevelopmental | 1.250 | 1.81 | 1.45 |
| Cesarean | ADHD | 1.388 | 2.12 | 1.42 |
| Cesarean | Speech/language | 1.200 | 1.69 | 1.19 |
| Preterm | Composite neurodevelopmental | 1.926 | 3.26 | 2.73 |
| Low birth weight | Composite neurodevelopmental | 2.640 | 4.72 | 3.79 |

### 8 Negative-Binomial Dispersion Diagnostics

Table S8: **Primary population negative-binomial shape estimates.** The table reports the fitted NB2 shape parameter $\theta$ from the primary population `MASS::glm.nb` model for each displayed outcome. Under the NB2 variance function, $\mathrm{Var}(Y) = \mu + \mu^2/\theta$; smaller $\theta$ therefore indicates stronger overdispersion relative to the mean. Intellectual disability is not shown because the population negative-binomial model did not converge at the theta boundary and the outcome was excluded from population modeling.

| **Outcome** | **Mean count** | **Zero, %** | **Shape $\theta$** |
|---|---|---|---|
| Composite neurodevelopmental | 2.937 | 85.5 | 0.0466 |
| Autism spectrum | 1.086 | 98.7 | 0.0031 |
| ADHD | 0.240 | 97.3 | 0.0153 |
| Conduct disorder | 0.027 | 99.1 | 0.0098 |
| Oppositional defiant disorder | 0.019 | 99.7 | 0.0017 |
| Anxiety disorders | 0.251 | 95.8 | 0.0260 |
| Obsessive-compulsive disorder | 0.013 | 99.8 | 0.0015 |
| Tic disorders | 0.010 | 99.5 | 0.0090 |
| Specific learning disorders | 0.025 | 99.6 | 0.0027 |
| Speech/language disorders | 1.322 | 91.2 | 0.0311 |
| Developmental coordination disorder | 0.102 | 98.5 | 0.0066 |
| Depressive disorders | 0.082 | 98.2 | 0.0149 |
| Elimination disorders | 0.031 | 98.6 | 0.0243 |

### 9 Cesarean Sensitivity Analyses

Table S9: **Cesarean sensitivity analyses.** Population rows are negative-binomial models with the same covariate set as the main arm. Sibling rows are negative-binomial fixed-effects estimates. Values are percent changes with 95% confidence intervals. The primary fitted support set starts from the constructed non-twin sibling cohort of 259,339 children in 123,926 families; model-specific zero-event birth-year exclusions remove 21 children and 9 family identifiers from the displayed primary cesarean fits, leaving 259,318 children in 123,917 families. The pregnancy-covariate-adjusted sibling row adds maternal-age indicators and pregnancy-varying maternal infectious, immune-mediated, and antimicrobial markers; it reports one fewer cesarean-discordant family (13,188 vs 13,189 in the primary row) under the expanded covariate set. These analyses were targeted robustness checks, not a new discovery matrix.

| Analysis | Restriction | N / families | Selected cesarean estimates |
|---|---|---|---|
| Population NB | Term, normal birth weight, no newborn antibacterial dispensing | $n = 929{,}236$ | Composite neurodevelopmental 23.8% (21.2, 26.5); speech/language 20.8% (17.7, 24.0); ADHD 11.2% (6.8, 15.9); autism spectrum 19.4% (10.1, 29.5). |
| Population NB | ≥5 years follow-up | $n = 258{,}078$ | Composite neurodevelopmental 23.2% (19.1, 27.4); speech/language 17.0% (12.0, 22.4); ADHD 14.0% (8.5, 19.7); autism spectrum 29.6% (15.3, 45.4). |
| Sibling NB-FE | Main analytic rows after model-specific zero-event-year exclusions | $n$ = 259,318; 123,917 families; 13,189 cesarean-discordant families | Composite neurodevelopmental 25.0% (10.6, 41.4); speech/language 20.0% (2.6, 40.3); ADHD 38.8% (9.5, 75.8); autism spectrum 5.0% (−42.6, 92.2). |
| Sibling NB-FE | Main analytic rows, pregnancy-covariate adjusted | $n$ = 259,318; 123,917 families; 13,188 cesarean-discordant families | Composite neurodevelopmental 24.8% (10.4, 41.1); speech/language 19.9% (2.5, 40.2); ADHD 37.0% (8.1, 73.5); autism spectrum 3.2% (−43.9, 89.7). |
| Sibling NB-FE | Term, normal birth weight, no newborn antibacterial dispensing | $n$ = 175,462; 84,315 families; 8,086 cesarean-discordant families | Composite neurodevelopmental 13.2% (−3.6, 32.9); speech/language 9.2% (−10.8, 33.9); ADHD **58.3**% (15.4, 117.2; $p = 0.0044$); autism spectrum −12.6% (−57.3, 78.9). |
| Sibling NB-FE | ≥5 years follow-up | $n$ = 59,023; 28,319 families; 2,743 cesarean-discordant families | Composite neurodevelopmental 26.7% (1.8, 57.7); speech/language 21.9% (−10.8, 66.7); ADHD 44.6% (2.6, 103.7); autism spectrum 93.8% (−44.5, 576.8). |

| Analysis | Restriction | N / families | Selected cesarean estimates |
|---|---|---|---|
| Low-risk population decomposition | Term, normal birth weight, no newborn antibacterial dispensing, no maternal peripartum bacterial/viral/other-infectious/immune-mediated diagnosis | $n$ = 804,231; claims-defined no-labor/no-repeat exposed $n$ = 74,303 | Claims-defined no-labor/no-repeat estimates: composite neurodevelopmental 48.0% (42.4, 53.7); speech/language 41.0% (34.5, 47.8); ADHD 24.1% (15.4, 33.6); autism spectrum 44.9% (25.1, 67.9). |

#### 10 Timing and Differential-Surveillance Checks

Table S10: **Timing and differential-surveillance checks.** These rows summarize the aggregate analyses used for manuscript timing-window and surveillance-probe claims.

| **Claim** | **Estimate or test** | **Analysis** |
|---|---|---|
| Antibacterial timing, composite neurodevelopmental | 32.5%, 24.7%, 6.0%; linear-trend $p = 5.3 \times 10^{-9}$ | Composite neurodevelopmental timing model and timing-interaction test tables |
| Antibacterial timing, ADHD | Mutually adjusted binary-window estimates 14.1%, 23.2%, 28.8%; separate earliest-window factor RRs 1.35, 1.41, 1.29 with empirical peak at 29–90 d, linear-component LRT $p = 8.9 \times 10^{-32}$ and deviation-from-linearity LRT $p = 4.17 \times 10^{-4}$ | ADHD timing model and timing-interaction test tables |
| Differential-surveillance injury floor | Injury RR 1.18 to 1.30 across preterm, low-birth-weight, and neonatal-antibacterial discordance | Differential-surveillance negative-control table |
| Well-child-adjusted shift | Preterm and low-birth-weight neurodevelopmental focal cells shift by at most 3.3 percentage points versus primary; low-birth-weight autism shifts by 12.9 percentage points versus primary | Well-child-visit sensitivity summary table |

#### 11 Data and Code Availability

Individual-level MarketScan data cannot be redistributed under the Merative license. Analysis code, figure source-data workbooks, and releasable aggregate summaries are maintained in the reviewer-access repository and will be released publicly where licensing permits. The submitted source-data workbooks support the displayed figures; additional aggregate model summaries support the supplementary tables and reproducibility checks.
